# Declining Pediatric Representation in NIH Artificial Intelligence and Machine Learning Funding, 2020–2024

**DOI:** 10.64898/2026.04.08.26350420

**Authors:** Vidith Phillips, Paravreet Woodwal

**Affiliations:** Department of Radiology, St. Jude Children’s Research Hospital, Memphis, Tennessee; School of Medicine, Johns Hopkins University, Baltimore, Maryland; Bloomberg School of Public Health, Johns Hopkins University, Baltimore, Maryland

## Abstract

**Background:** Artificial intelligence and machine learning (AI/ML) are among the fastest-growing domains in NIH research funding, but whether children have shared equitably in this expansion is unknown. We characterized pediatric representation in NIH AI/ML funding from fiscal years (FY) 2020 to 2024.

**Methods:** NIH grant data were obtained from Research Portfolio Online Reporting Tools Expenditures and Results bulk files for FY2020 to FY2024. AI/ML grants were identified using the NIH Research, Condition, and Disease Categorization “Machine Learning and Artificial Intelligence” category, and pediatric grants using the “Pediatric” category. Subprojects were excluded. Grants were deduplicated within each fiscal year by core project number for trend analyses and across all years retaining the most recent fiscal year for cross-sectional totals. Disease areas were identified by keyword searches of titles and abstracts.

**Results:** Across FY2020 to FY2024, 5,624 unique NIH AI/ML grants totaling $3,371 million were identified. Of these, 836 grants (14.9%) were classified as pediatric, representing $401 million (11.9%) of total NIH AI/ML funding. Although this share was consistent with the historically reported overall NIH pediatric funding baseline of approximately 10% to 12%, it remained substantially below the US pediatric population share of approximately 22%. The pediatric share of NIH AI/ML funding declined from 12.3% in FY2020 to 10.8% in FY2024, despite growth in absolute pediatric funding. Indexed to FY2020, pediatric AI/ML funding grew approximately 2.6-fold compared with 3.0-fold growth in the total portfolio. Across disease areas, unadjusted adult/general-to-pediatric funding ratios ranged from 2.0-fold in mental health to 9.8-fold in cancer.

**Conclusions:** Pediatric representation in NIH AI/ML funding remained low and declined over time as the overall portfolio expanded. These findings suggest that growth in NIH AI/ML investment has not been matched by proportional gains for pediatric research.

## Introduction

Artificial intelligence and machine learning (AI/ML) represent the fastest-growing domain in NIH research funding, yet whether children have shared equitably in this expansion remains uncharacterized. Pediatric research has historically received approximately 10–12% of the NIH budget, a share that correlates poorly with pediatric disease burden^1^ and raises concern that structural underinvestment may compound as AI reshapes biomedical research priorities. A September 2025 executive order directing NIH to harness AI to unlock cures for pediatric cancer^2^ sharpens this concern and underscores the need for a rigorous empirical baseline. Although a recent cross-sectional analysis documented rapid NIH AI/ML portfolio growth from FY2019 to FY2023, pediatric representation was not examined.^3^ This study aimed to characterize NIH pediatric AI/ML funding from FY2020 to FY2024 and benchmark it against the overall NIH pediatric funding baseline and US population share.

## Methods

NIH grant data were obtained from the Research Portfolio Online Reporting Tools Expenditures and Results (RePORTER) bulk download files for FY2020 to FY2024.^4,5^ AI/ML grants were identified using the NIH Research, Condition, and Disease Categorization (RCDC) “Machine Learning and Artificial Intelligence” spending category; pediatric grants were identified using the RCDC “Pediatric” spending category. Subprojects were excluded. Grants were deduplicated within each fiscal year by core project number for trend analyses, and across all years retaining the most recent fiscal year for cross-sectional totals. FY2025 was excluded due to unavailability of RCDC assignments.

Disease areas were identified through keyword searches of grant titles and abstracts; categories were non-exclusive. This study used publicly available deidentified data, was exempt from institutional review board oversight, and adhered to Strengthening the Reporting of Observational Studies in Epidemiology (STROBE) reporting guidelines.

## Results

Across FY2020 to FY2024, we identified 5,624 unique NIH AI/ML grants totaling $3,371 million. Of these, 836 grants (14.9%) were classified as pediatric, representing $401 million (11.9%) of total NIH AI/ML funding (**Table1, Fig.1A**). This 11.9% share is consistent with the historically reported 10–12% overall NIH pediatric funding baseline,^1^ yet remains substantially below the US pediatric population share of approximately 22%^6^ (**Fig.1A**). Ninety-three pediatric grants (11.1%), totaling $51 million, were administered by children’s hospitals.

**Table 1.**
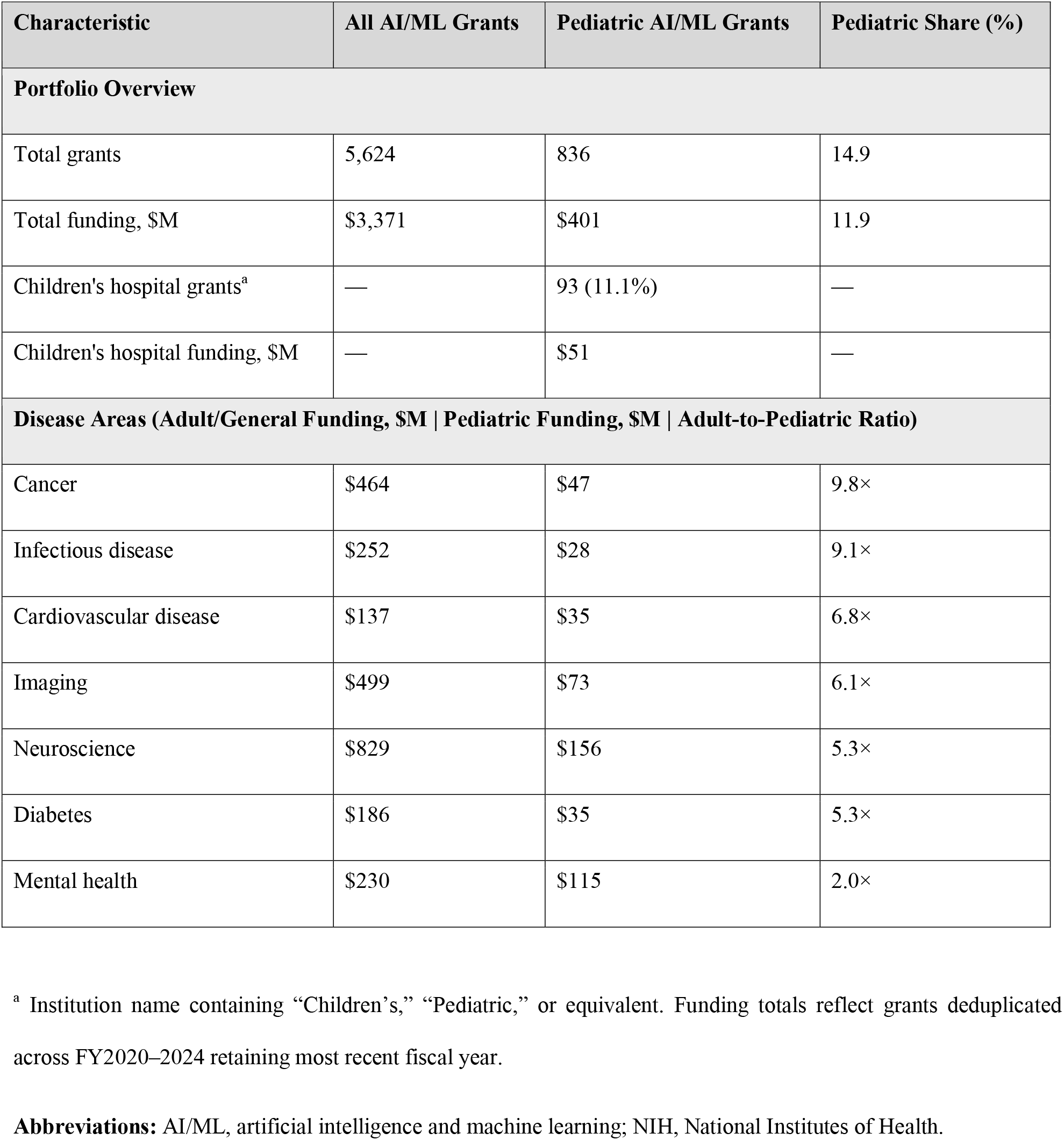
Characteristics of NIH-Funded Artificial Intelligence and Machine Learning Research, Fiscal Years 2020 to 2024.

**Figure 1.**
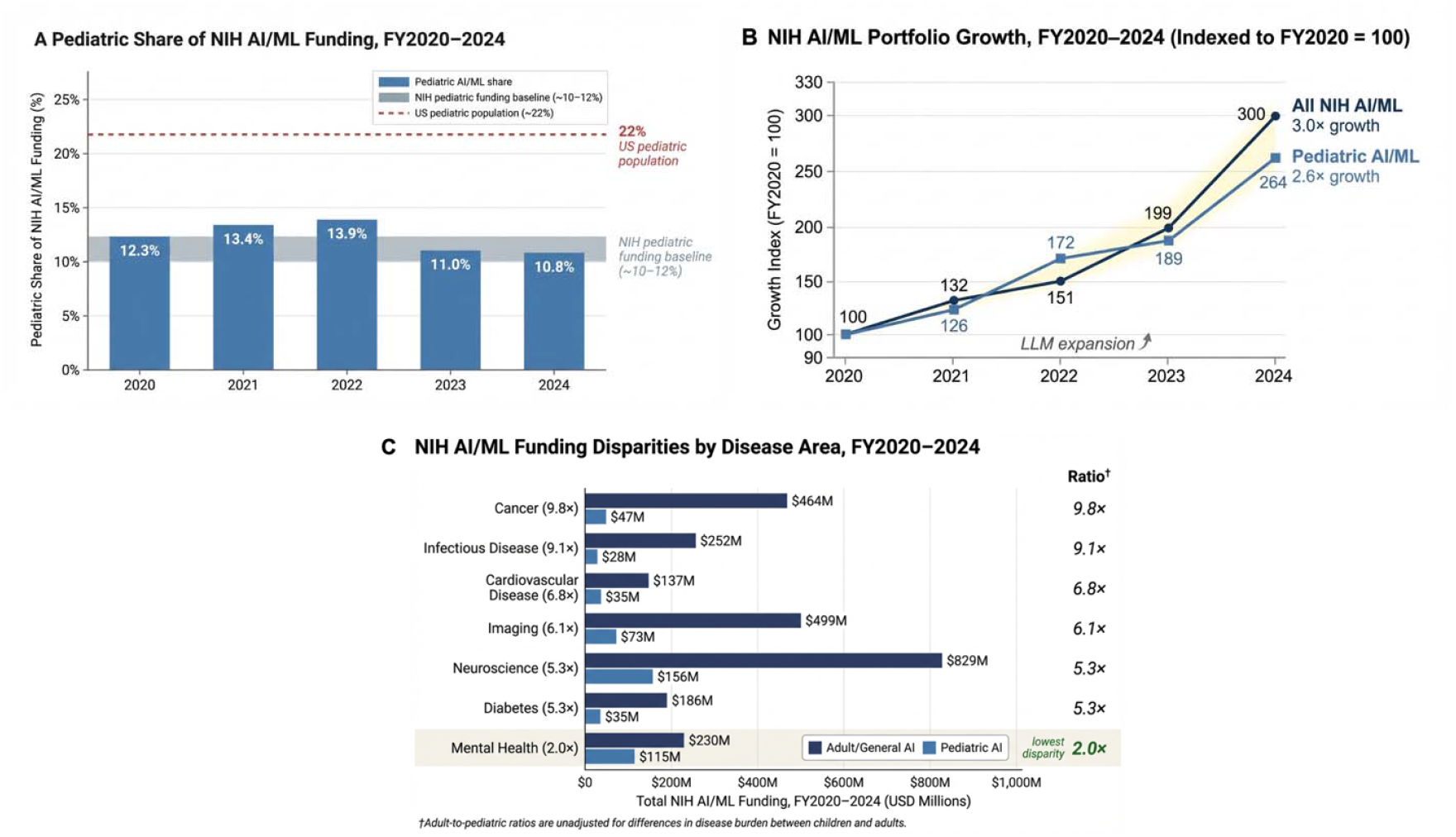
NIH Pediatric AI/ML Funding, FY2020–2024. (**A**) Pediatric AI/ML grants as a share of total NIH AI/ML funding by fiscal year. The shaded band represents the overall NIH pediatric research funding baseline (∼10–12%); the dashed line represents the US pediatric population share (∼22%). (**B**) Total versus pediatric NIH AI/ML funding growth, indexed to FY2020 = 100. Pediatric AI/ML funding grew 2.6-fold compared with 3.0-fold growth in the total portfolio; divergence accelerated from FY2022 onward. (**C**) Adult/general and pediatric NIH AI/ML funding .across seven disease areas, sorted by adult-to-pediatric funding ratio. Ratios ranged from 9.8× (cancer) to 2.0× (mental health). †Adult-to-pediatric ratios are unadjusted for differences in disease burden between children an adults. Grants were identified using NIH RePORTER RCDC spending categories “Machine Learning and Artificial Intelligence” and “Pediatric.” FY2025 excluded due to unavailability of RCDC assignments. AI/ML indicates artificial intelligence and machine learning; NIH, National Institutes of Health; RCDC, Research, Condition, an Disease Categorization.

Despite growth in absolute pediatric AI/ML funding over the study period, the pediatric share declined from 12.3% in FY2020 to 10.8% in FY2024 (**Fig.1B**). Indexed to FY2020, pediatric AI/ML funding grew approximately 2.6-fold compared with 3.0-fold growth in the total portfolio, with divergence accelerating from FY2022 onward.

Unadjusted adult-to-pediatric funding ratios varied markedly across disease areas and should be interpreted as descriptive baselines rather than equity measures, given differences in pediatric versus adult disease burden across conditions (**Fig.1C**). The largest ratios were observed in cancer (9.8×; $47M pediatric vs $464M adult/general) and infectious disease (9.1×; $28M vs $252M); imaging, cardiovascular disease, neuroscience, and diabetes ranged from 5.3× to 6.8×. Mental health showed the smallest gap (2.0×; $115M pediatric), possibly reflecting longstanding National Institute of Mental Health investment in pediatric behavioral research.

## Discussion

The declining pediatric share of NIH AI/ML funding signals a structural disadvantage: as AI/ML rewards computational scale, pediatric research is systematically less able to compete given its reliance on smaller and more heterogeneous datasets. That the 11.9% overall share remains consistent with the historical NIH pediatric baseline offers little reassurance; prior work demonstrated that this baseline itself correlates poorly with pediatric disease burden,^1^ and benchmarking AI/ML equity against an inadequate floor risks normalizing a gap that is widening. The consequences extend into clinical translation: only 17% of FDA-authorized AI/ML devices are labeled for pediatric use, fewer than one in five were validated using pediatric data,^7^ and fewer than 3% of FDA-cleared AI imaging tools indicate pediatric use.^8^ Study limitations include restriction to NIH funding only, exclusion of FY2025 data, use of population share as an imperfect equity benchmark, and reliance on keyword-based disease classification with non-exclusive categories.

Correcting this trajectory requires structural intervention, not incremental accommodation. Dedicated NIH funding mechanisms for pediatric AI, mandatory reporting of pediatric validation in grant outcomes, disease-burden-weighted equity benchmarks, and restoration of RCDC categorical spending transparency for FY2025 and beyond would each address a distinct failure point. Without these, children risk becoming the population that AI in medicine was never built to serve.

## Data Availability

Deidentified individual participant data will not be made available. All underlying data are publicly accessible through NIH RePORTER at https://reporter.nih.gov

## Abbreviations

AI/ML: artificial intelligence and machine learning
NIH: National Institutes of Health
RCDC: Research, Condition, and Disease Categorization
RePORTER: Research Portfolio Online Reporting Tools Expenditures and Results.

